# Evaluating the effects of Toronto’s supervised consumption sites on local crime

**DOI:** 10.1101/2024.11.08.24316990

**Authors:** Dimitra Panagiotoglou, Jihoon Lim, Geoff Ingram, Mariam El Sheikh, Imen Farhat, Xander Bjornsson, Maximilian Schaefer

**Affiliations:** Department of Epidemiology, Biostatistics and Occupational Health, McGill University, Montreal, Québec, Canada

**Author notes:** Corresponding Author Dimitra Panagiotoglou, PhD 2001 McGill College Avenue Montreal, QC H3A 1G1.

**Keywords:** harm reduction, supervised consumption sites, public health, crime, neighbourhoods

## Abstract

**Importance:** Beginning in August 2017, nine overdose prevention sites and supervised consumption sites (OPS/SCS) began operating in Toronto, Canada. Following years of community pushback that these sites increased local crime and disorder, they were closed in March 2025.

**Objective:** To evaluate the population-level effects of OPS/SCS on crime and disorder.

**Design, Setting, Participants:** This two-part ecological study used Toronto Police Service data to compare crime incidence before and after OPS/SCS implementation using interrupted time series analyses with and without controls. We restricted analysis to crimes that occurred within city boundaries between 1 January 2014 and 30 June 2024.

**Main Outcomes and Measures:** We used monthly event counts of all assaults, auto thefts, break and enters, robberies, thefts over $5000, bicycle thefts, thefts from motor vehicle and mental health apprehensions as our eight outcomes. We compared incidence within 100m, 200m, and 400m of the geolocation of each OPS/SCS before and after implementation; and repeated analysis using treated and synthetic control neighbourhoods. We pooled estimates for population-level effects.

**Results:** Within 400m (approximately a quarter mile), we observed level effects for break and enters (48.87%, 95% CI: 26.15, 75.68%) and bicycle thefts (-19.46%, 95% CI: -30.36, -6.86%) immediately post-implementation. Meanwhile, monthly trends for assaults (-0.5%, 95% CI: -0.92, -0.07%), break and enters (-1.11%, 95% CI: -1.60, -0.61%), robberies (-1.36, 95% CI: -1.96, -0.75%), thefts over $5000 (-1.47%, 95% CI: -2.92, 0.01%), bicycle thefts (-1.54%, 95% CI: -2.44, -0.62%), and thefts from vehicles (-1.59%, 95% CI: -2.58, -0.60%) declined. Pooled neighbourhood analyses showed level-effects for break and enters (18.83%, 95% CI: 1.70, 38.84%) and mental health apprehensions (10.20%, 95% CI: 1.55, 19.60%) post-implementation; and monthly changes in trends for break and enters (-0.46%, 95% CI: - 0.93, 0.01%), auto theft (-0.66%, 95% CI: -1.31, 0.00%), thefts over $5000 (-1.02%, 95% CI: -2.05, 0.03%), and bicycle thefts (-1.07%, 95% CI: -1.58, -0.57%). Site- and neighbourhood-specific results revealed some communities were impacted while others were not.

**Conclusions and Relevance:** The effects of Toronto’s OPS/SCS on crime were generally neutral. Break and enters increased immediately post-implementation, but declined with time.

**Data Sharing Agreement:** This study used publicly available data provided by Toronto Police Services. For more information, please visit the Toronto Police Service Public Safety Data Portal at: https://data.torontopolice.on.ca/. For Statistics Canada census tract profiles visit: https://www12.statcan.gc.ca/census-recensement/2021/dp-pd/prof/index.cfm?Lang=E.

**Key points:** **Question:** What was the effect of overdose prevention sites and supervised consumption sites (OPS/SCS) on crime in Toronto?

**Findings:** Pooled site- and neighbourhood-level random effects models found counts of break and enters increased, but trends in outcomes declined per month. Trends in assaults, robberies, thefts over $5000, bicycle thefts, and thefts from motor vehicles also decreased. Site- and neighbourhood-specific interrupted time series showed some communities were impacted negatively while others were not.

**Meaning:** The effects of Toronto’s OPS/SCS on crime were generally neutral to positive; break and enters increased immediately post-implementation, but declined with time.

## Introduction

Canada’s largest city, Toronto, has been hit hard by the ongoing opioid crisis. Since 2016 there have been over 3,251 opioid-related deaths; and in 2021 the death rate exceeded 19.4 deaths per 100,000,^1^ putting it on par with cities like New York, Houston, San Diego, Las Vegas, and Seattle.^2^ As part of the city’s harm reduction strategy, nine overdose prevention sites (OPS) and supervised consumption sites (SCS) were implemented between 2017 and 2018. Despite evidence demonstrating the health benefits of OPS/SCS for people who use unregulated drugs,^3,4^ these sites remain controversial. Critics argue OPS/SCS may increase local crime by attracting drug-related activity such as theft, assault and open drug use.^5^

Early evidence following the implementation of Vancouver’s Insite (2003), and Sydney’s Medical Injecting Centre (2001) found no changes in police-recorded thefts or robberies, drug possession, drug dealing, open drug use or assaults.^8–10^ More recently, a study examining the effects of an unsanctioned SCS in the United States reported assault, burglary, larceny theft, robbery and drug related incidents declined in the treated neighbourhood but remained the same in two control communities.^11^ Similarly, a study examing the effects of New York’s two overdose prevention sites observed no significant effects on counts of violent or property crimes, in aggregate.^12^ However, the study did reveal a 30.4% (95%CI: 10.4, 54.0%) increase in aggravated assaults, and a 69.1% (95%CI: 18.3, 141.7%) increase in vehicle thefts offset by the 34.9% (95%CI: -54.0, -7.8%) decline in robberies. Meanwhile, a second study of New York’s sites found differential effects of the SCS owing to plausible moderating factors including a ‘target of opportunity’ for petty larceny near one site but not the other.^13^

While many studies indicate SCS do not increase neighbourhood crime and may reduce public drug use, differences in the methods used and their respective limitations may explain the variations in outcomes reported. These include short observation periods and the inability to detect meaningful differences in crime trends over time, aggregating crimes to improve statistical power but losing nuance, and not adjusting for local population demographics. Further, many of the studies dominating the literature are specific to well-resourced sites and, in the case of Vancouver’s Insite, set up as a pilot to demonstrate feasibility and impact.^14^ Given the ongoing equipoise, and local arguments opposed to the continued operation of Toronto’s sites, we evaluated the effects of the city’s nine OPS/SCS on neighbourhood crime and mental health act apprehensions using publicly available data.

## Method

### Study design

For this ecological study, we used interrupted time series analyses with and without synthetic controls.

### Setting

Toronto is an ethno-racially diverse city with approximately 2.8 million residents. ^15,16^ Between 1990 and 2022, the city was divided into 140 neighbourhoods with unique demographics and social service needs. Each neighbourhood is comprised of multiple, contiguous Statistics Canada census tracts.^17^ Although some neighbourhoods were redefined to reflect demographic and density changes in spring 2022, we refer to the original 140 neighbourhood boundaries in our study. Beginning in 2017, the majority of opioid poisoning calls were observed to be concentrated in downtown neighbourhoods.^18^ In response, the Overdose Prevention Society opened the first unsanctioned OPS in Moss Park in August 2017 and within a year, nine OPS/SCS were in operation (Supplement Table 1).

### Data

We restricted analysis to founded crimes that occurred between 1 January 2014 and 30 June 2024 as reported in four publicly available Toronto Police Service datasets: Major Crime Indicators (MCI), Bicycle Thefts, Theft from Motor Vehicle, and Mental Health Act Apprehensions (Supplement Table 2 for definitions). The MCI included records for five major crimes: assaults, auto thefts, break and enters, robberies, and thefts over $5000. Across all four datasets, each incident record included a unique identifier, date of occurrence, and the name and numeric identifier of the neighbourhood. Apart from mental health act apprehensions, the remaining three datasets also included geo-coordinates of the intersection closest to the event location.

### Analysis

We conducted two sets of analysis to overcome the limitations observed in other studies. First, we conducted site-specific interrupted time series (ITS) analysis with segmented negative binomial regression to test for level and trend changes on counts of crimes within 100m, 200m and 400m (i.e., approximately a quarter mile) of the nine OPS/SCS: *outcome_jt_* =β_0_ +β_1_ ⋅ *time_t_* +β_2_ ⋅ *level_j_* +β_3_ ⋅ *trend_jt_* +ε*_jt_,* where β_1_ is the underlying trend in the outcome across time, β_2_ captures the level effect immediately post-implementation, and β_3_represents the trend effect following the implementation of the local OPS/SCS relative to the underlying trend. To avoid double counting events that occurred within the radii of observation of two sites (e.g., Fred Victor site opened within 400m of Moss Park OPS) we only assigned events that occurred outside the first site’s radius to the new site. We set the month of implementation as time zero, and reported monthly sums of outcomes.

We repeated the analysis comparing outcome counts in treated neighbourhoods with synthetic controls to account for neighbourhood-level demographic differences. We linked Toronto Police Service records with 2011, 2016, and 2021 census tract profiles using their respective year of incidence and boundary files for point-in-polygon spatial joins, and created neighbourhood demographic profiles by weighting the census tracts that fell within each neighbourhood’s boundaries.^19^ For each, treated neighbourhood we made a synthetic control neighbourhood using the pool of neighbourhoods that never implemented an OPS/SCS during our observation period and whose boundaries were > 400m of any OPS/SCS.^20^ In creating synthetic controls, we accounted for population size and density, proportion of females, distribution of age (0-14, 15-64, and 65+) and average age, average household size, median income, prevalence of low-income households after tax, unemployment rate, and proportion of lone-parent households, immigrants, visible minorities, and individuals without high school diploma or equivalent certificate.

We used controlled interrupted time series analysis with segmented negative binomial regression to compare the level and trends of crime and mental health act apprehension counts observed in treated neighbourhoods vs. synthetic controls post-implementation: *outcome_jkt_* =β_0_ +β_1_ ⋅ *time_t_* +β_2_ ⋅ *group_k_* + β_3_ ⋅ *group_k_* ⋅ *time_t_* +β_4_ ⋅ *level_jt_*+ β_5_ ⋅ *trend_jt_* +β_6_ ⋅ *level_jt_* ⋅ *group_k_* + β_7_ ⋅ *trend_jt_*⋅ *group_k_* +ε *_jkt_* where β_1_ is the underlying trend in the outcome observed in the synthetic control group, β_2_ is the level difference pre-intervention between the treated and synthetic control group, β_3_ represents the difference in trend between the treated and synthetic control group *before* the intervention is implemented, β_4_ is the level change in the synthetic control group post-implementation compared with immediately before the intervention’s implementation, β_5_ is the trend change for the synthetic control post-implementation, β_6_ is the level change of the treated group *relative* to the synthetic control group change observed post-implementation, and β_7_ is the trend change in the treated group *relative* to the synthetic control group post-implementation.

For both sets of models, we included four harmonic terms to capture seasonal effects, and used Newey-West standard errors with lag of 3 to account for data heteroscedasticity and residual autocorrelation. For overall effects, we pooled site- and neighbourhood-specific ITS outcomes and reported random effect estimates owing to the varying between-site heterogeneity.

All data preparation and analyses were conducted using R version 4.3.1 in R studio, with packages geosphere, AICcmodavg, tidyverse, sf, Synth, crsuggest, cancensus, foreign, tsModel, lmtest, Epi, splines, vcd, sandwich, reshape2, SCtools, janitor, gdata, rlang, tidyselect, car, and nlme.

### Ethics

This study used publicly available data and was exempt from ethics review by McGill’s institutional review board.

## RESULTS

After removing duplicates, errors, and incidents that occurred outside city boundaries, 172,681 assaults, 56,414 auto thefts, 71,255 break and enters, 26,180 robberies, 12,528 thefts over $5000, 29,128 bicycle thefts, 92,803 thefts from motor vehicles, and 110,387 mental health act apprehensions occurred within Toronto during the observation period. Of these, 21,705 (12.6%) assaults, 1,805 (3.2%) auto thefts, 6,808 (9.6%) break and enters, 4,103 (15.7%) robberies, 1,085 (8.7%) thefts over $5000, 5,586 (19.2%) bicycle thefts, and 8,286 (8.9%) thefts from motor vehicles occurred within 400m of the OPS/SCS locations.

Figure 1 summarizes the results of our site- (400m) and neighbourhood-specific analyses. For additional information on sites, their implementation dates, and locations; along with the creation of synthetic controls and results at 100m and 200m see Supplement.

**Figure 1.**
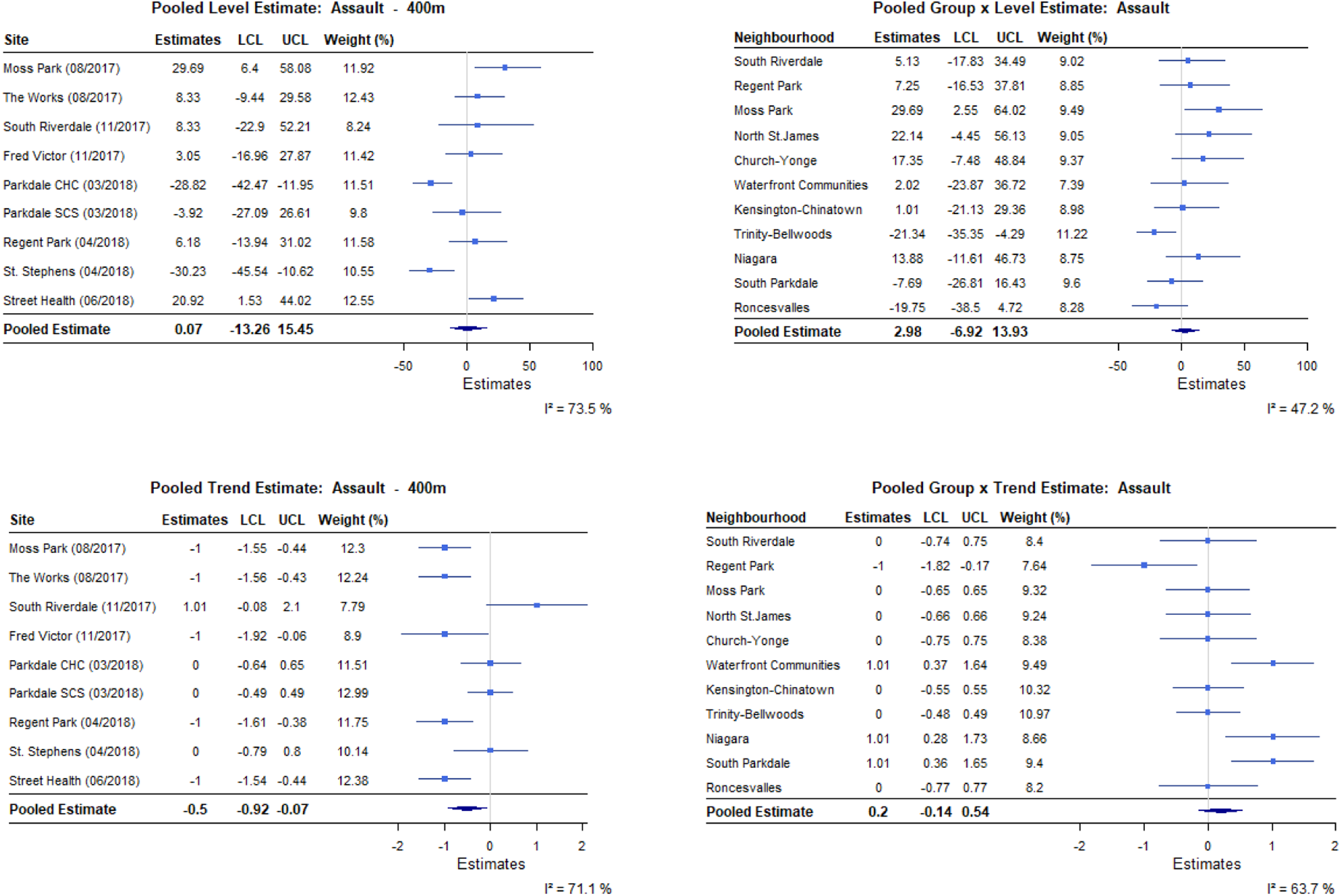
a. Pooled level and trend percent effect estimates within 400m of OPS/SCS and for treated compared with synthetic neighbourhoods, assaults

**Figure 1.**
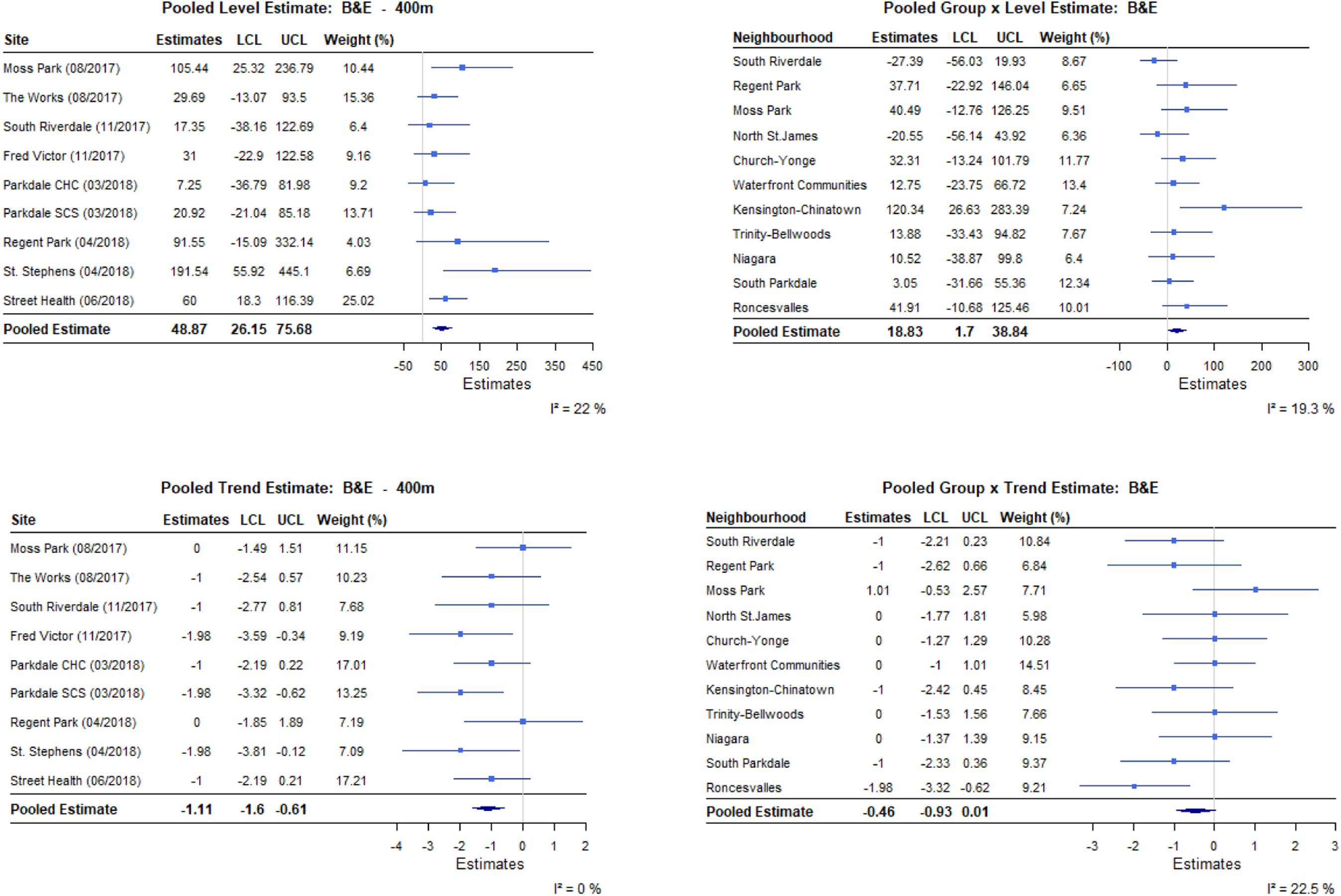
b. Pooled level and trend percent effect estimates within 400m of OPS/SCS and for treated compared with synthetic neighbourhoods, break and enters

**Figure 1.**
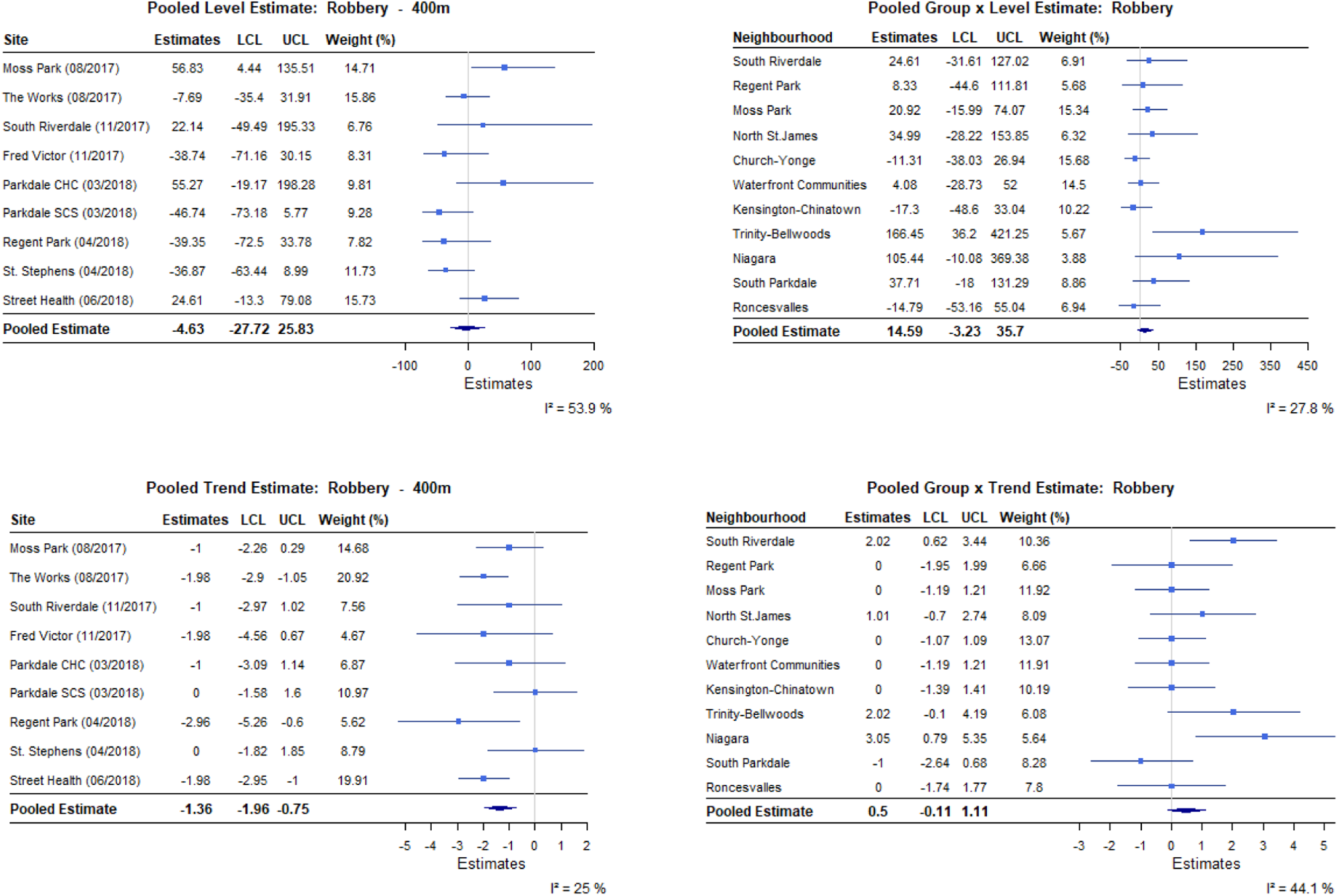
c. Pooled level and trend percent effect estimates within 400m of OPS/SCS and for treated compared with synthetic neighbourhoods, robberies

**Figure 1.**
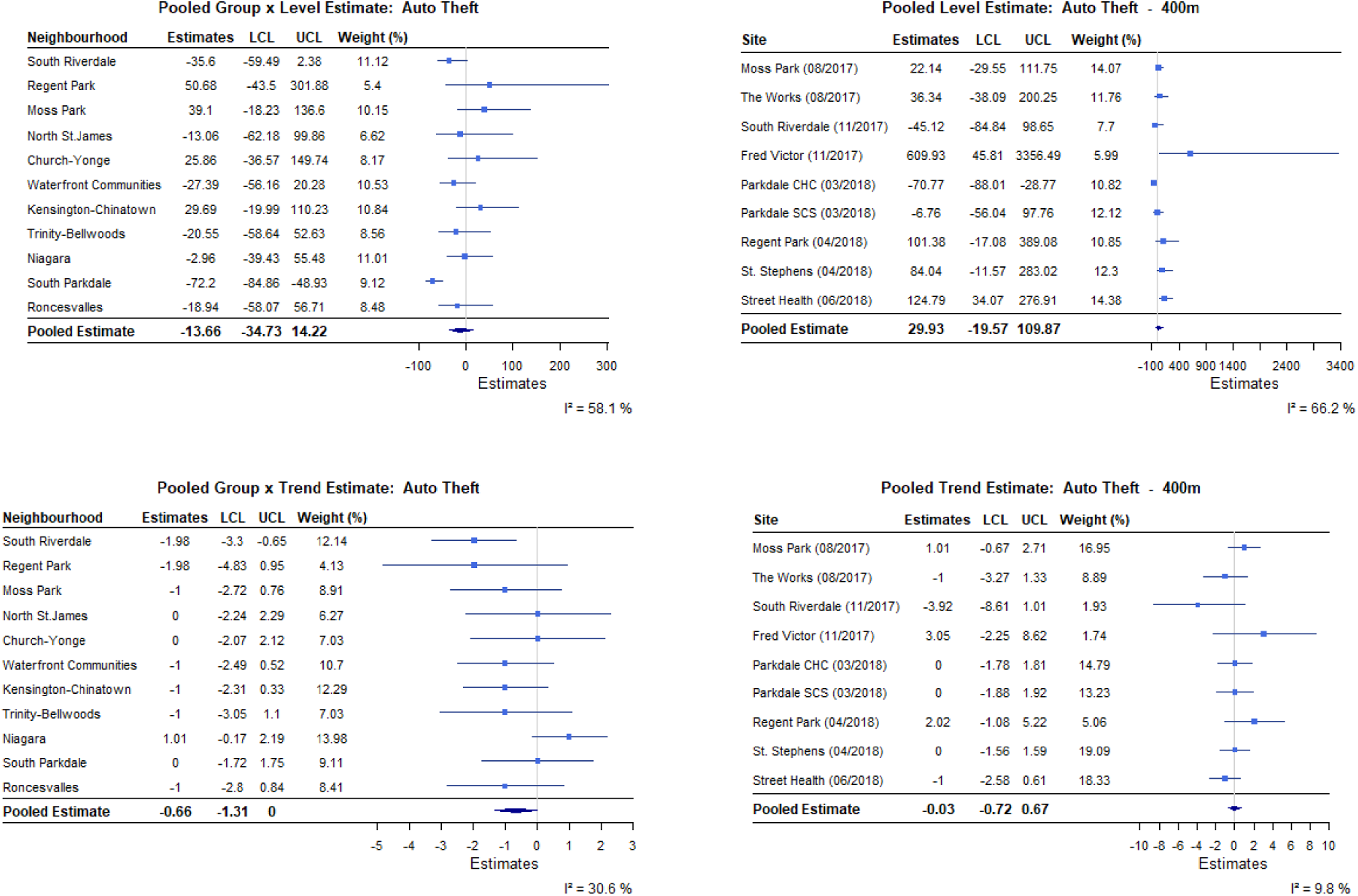
d. Pooled level and trend percent effect estimates within 400m of OPS/SCS and for treated compared with synthetic neighbourhoods, auto theft

**Figure 1.**
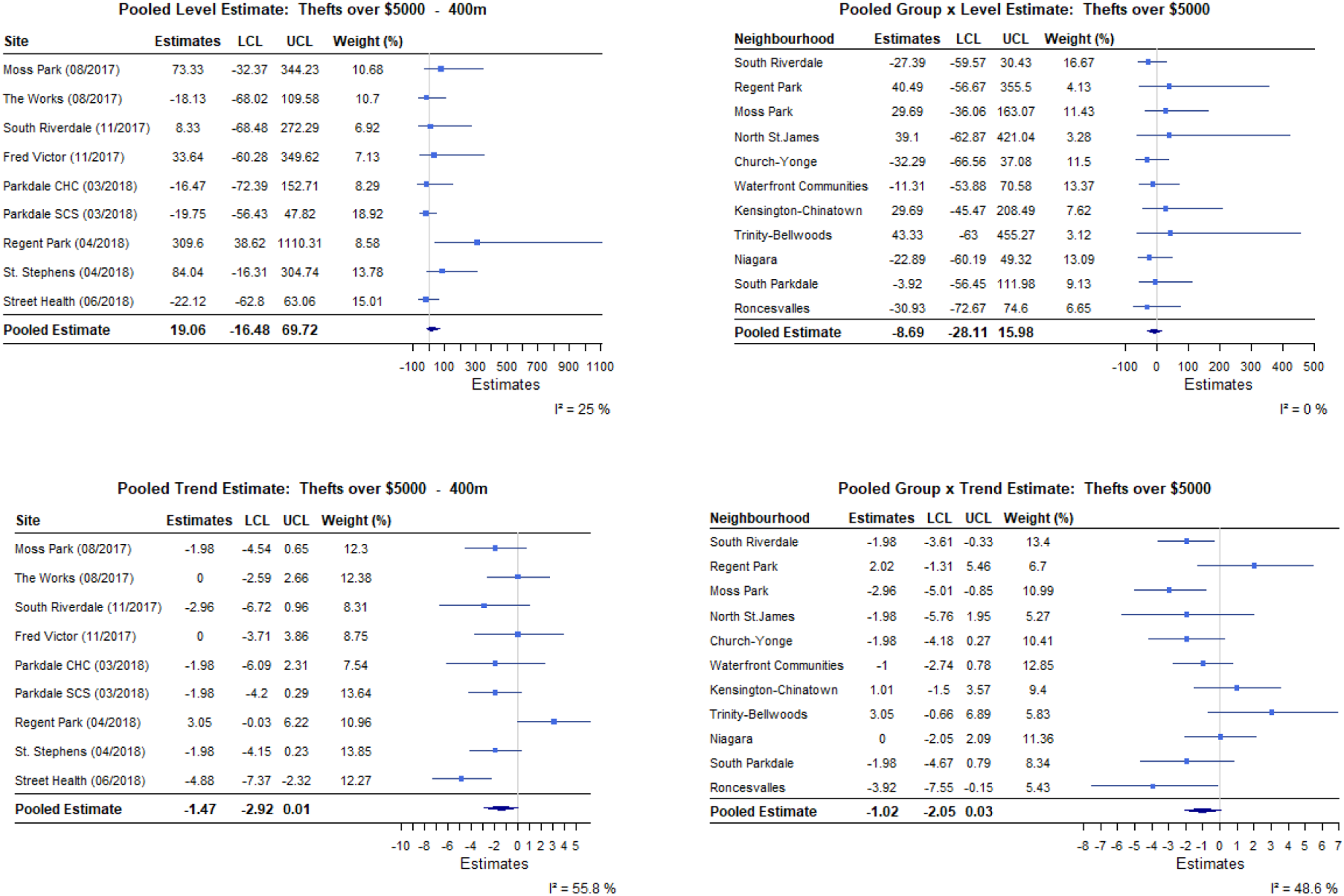
e. Pooled level and trend percent effect estimates within 400m of OPS/SCS and for treated compared with synthetic neighbourhoods, thefts >$5000

**Figure 1.**
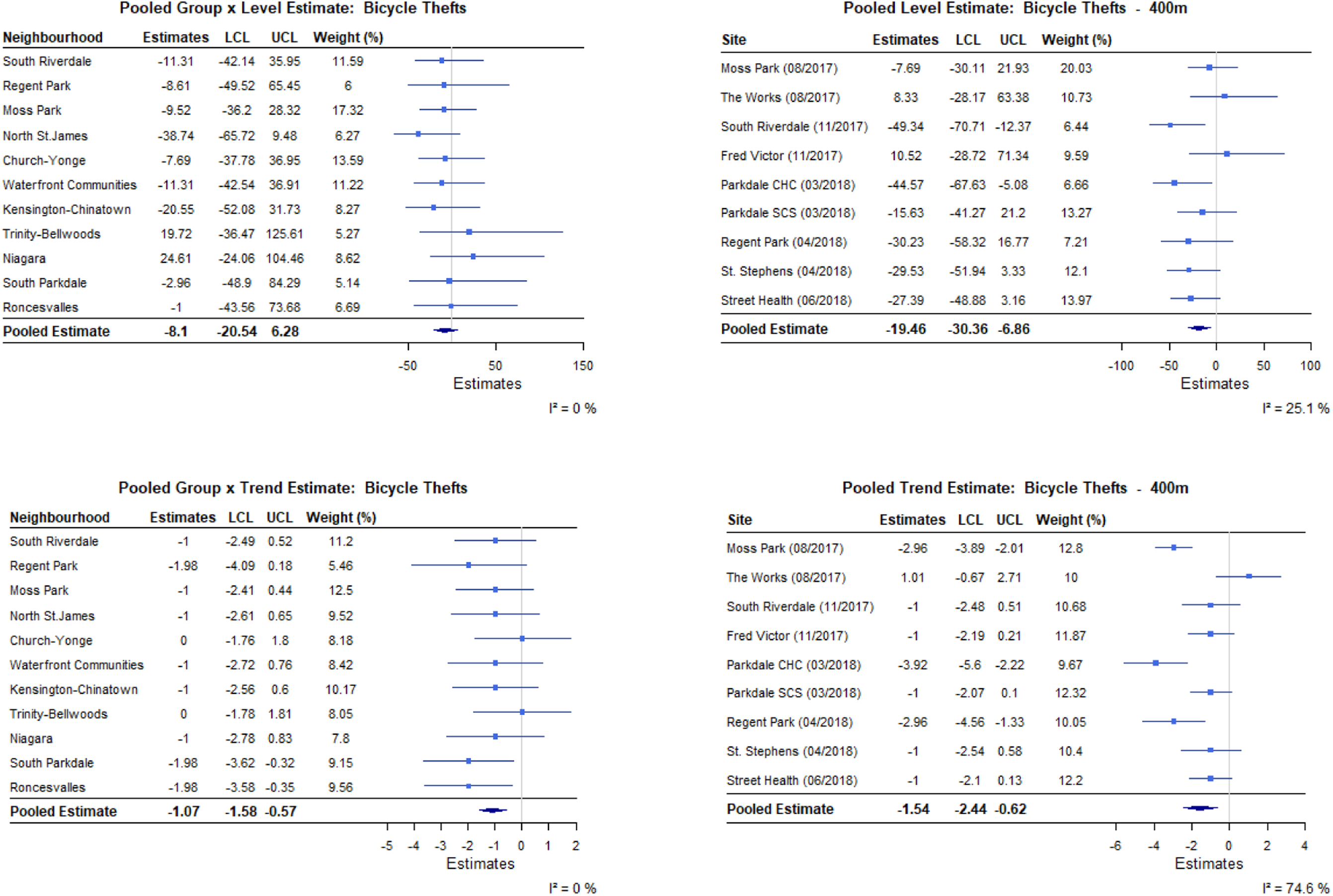
f. Pooled level and trend percent effect estimates within 400m of OPS/SCS and for treated compared with synthetic neighbourhoods, bicycle thefts

**Figure 1.**
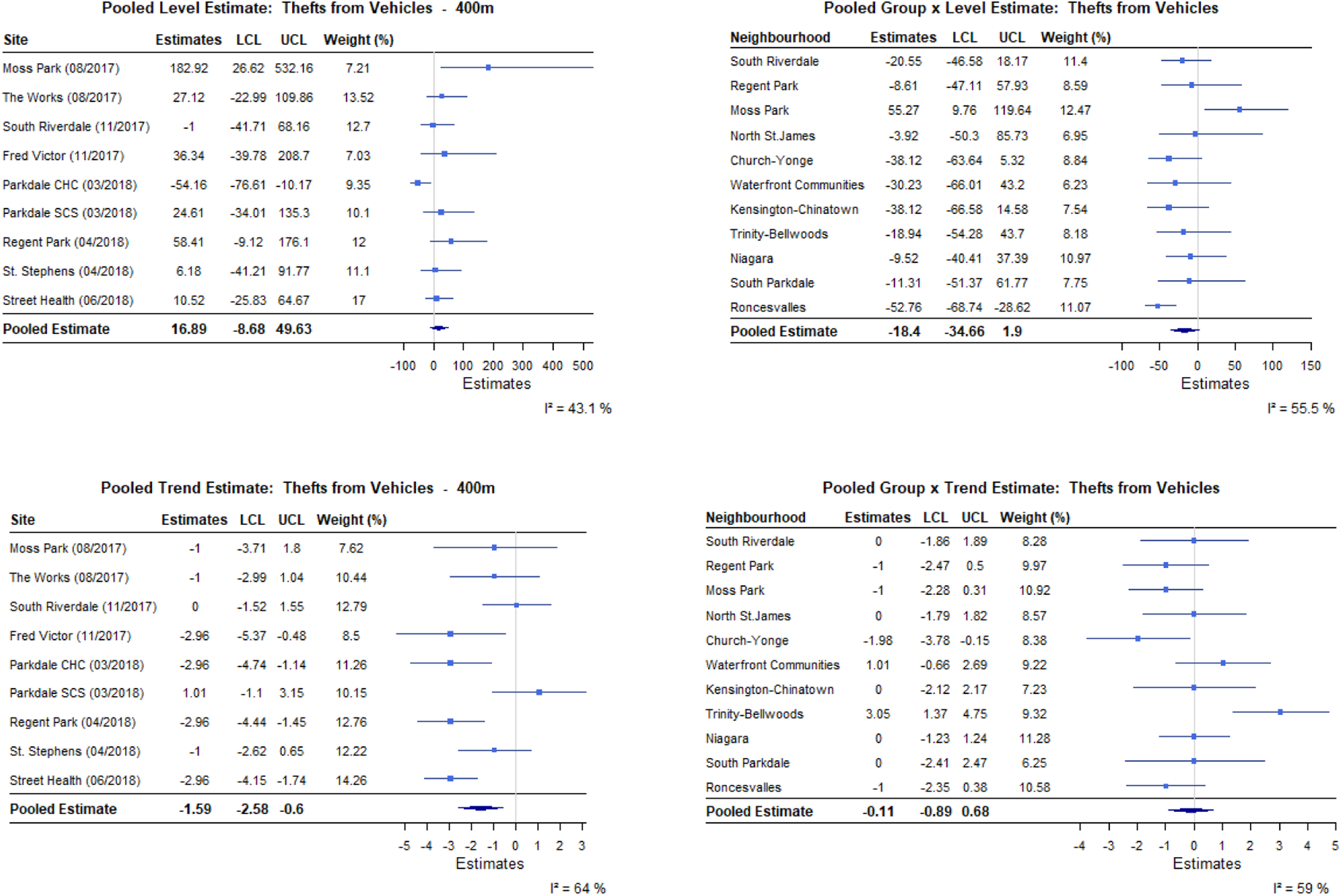
g. Pooled level and trend percent effect estimates within 400m of OPS/SCS and for treated compared with synthetic neighbourhoods, thefts from vehicles

**Figure 1.**
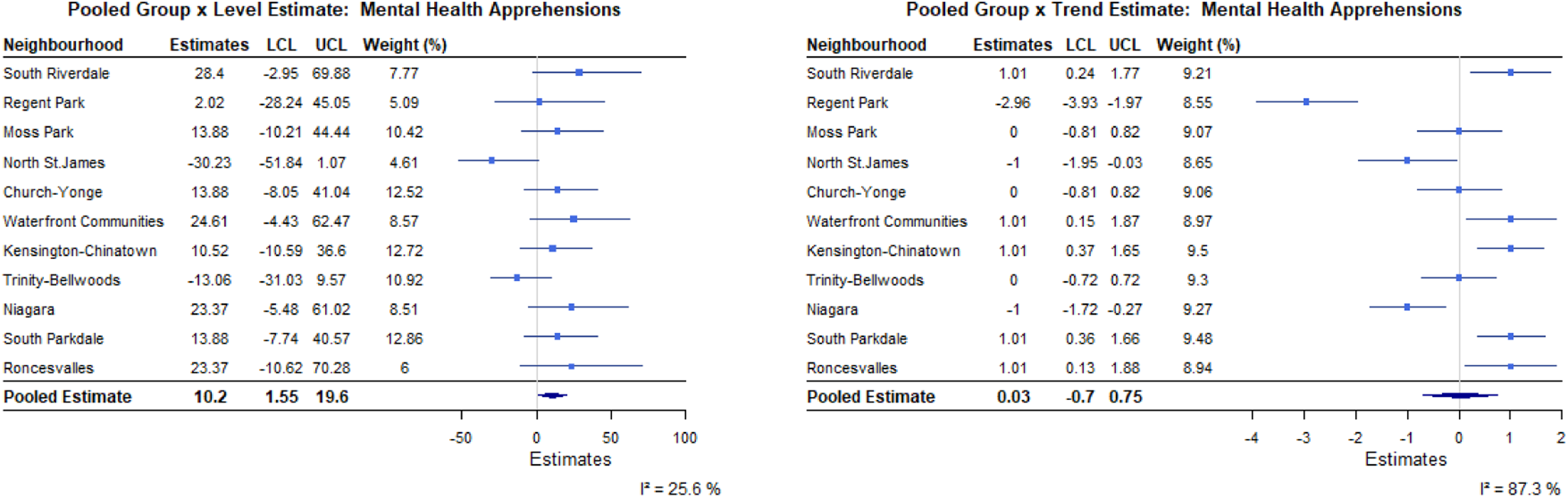
h. Pooled level and trend percent effect estimates for treated compared with synthetic neighbourhoods, mental health apprehensions

In pooled analysis, within 100m of OPS/SCS, there were no statistically significant level or trend effects in incidents of assaults, robberies, auto thefts, bicycle thefts, or thefts from motor vehicles. There were no level effects for break and enters, and thefts over $5000; but monthly trends declined 2.61% (95%CI: - 4.49, -0.68%) and 3.57% (95%CI: -6.87, -0.14%), respectively.

Within 200m, the lack of observed level or trend effects for assaults, auto thefts and thefts over $5000 persisted. There were no level effects observed for robberies, bicycle thefts, and thefts from motor vehicles; but trends declined 1.76% (95%CI: -2.54, -0.97%), 1.58% (95%CI: -2.71, -0.44%), and 1.60% (95%CI: -2.89, -0.29%), respectively. Meanwhile, break and enters increased 63.59% (95%CI: 9.23, 145%) immediately post-implementation, while trends declined 1.3% (95CI: -2.05, -0.53%) per month.

Within 400m, the level and trend effects observed within 200m of sites persisted for break and enters, robberies, auto thefts, and thefts from motor vehicles. Trends declined for assaults 0.5% (95%CI: -0.92, - 0.07%), and thefts over $5000 1.47% (95%CI: -2.92, 0.01). Bicycle thefts decrease 19.46% (95%CI: -30.36, -6.86%) immediately post-implementation, and continued to decline 1.54% (95%CI: -2.44, - 0.62%) each month thereafter.

Neighbourhood analyses found no level or trend effects for assaults, robberies, or thefts from motor vehicles. Break and enters increased 18.83% (95%CI: 1.70, 38.84%) immediately post-implementation but declined 0.46% (95%CI: -0.93, 0.01%) month-to-month. Trends declined for auto thefts 0.66% (95%CI: -1.31, 0.00%), thefts over $5000 1.02% (95%CI: -2.05, 0.03%), and bicycle thefts 1.07% (95%CI: -1.58, -0.57%). Meanwhile, we observed a level increase in mental health act apprehensions (10.20%, 95%CI: 1.55, 19.60%), with no trend effects.

Disaggregated results revealed differences in level and trend effects by site and neighbourhood. For example, assaults within 400m of Moss Park OPS increased (i.e., level effect) post-implementation, but decreased for both Parkdale Community Health Centre (CHC) and St. Stephen’s sites. Meanwhile, break and enters increased near The Works, Regent Park, and St. Stephen’s sites. At the neighbourhood level, assault trends increased for South Parkdale, Niagara, and Waterfront Communities, but declined for Regent Park. A level increase in break and enters was only observed for Kensington-Chinatown, while robbery effects were observed in Trinity-Bellwoods.

## DISCUSSION

Our analysis revealed that OPS/SCS did not have a level or trend effect on counts of grand theft or crimes against persons; and a protective effect on petty theft within 400m. The observed increase in break and enters within 400m diminished with time. A lack of effect on grand theft may reflect the absence or non-differential presence of organized criminal activity near OPS/SCS.^21^ Grand theft relies on concerted criminal networks targeting high value items; and is less sensitive to small sociodemographic changes at the community-level such as increases in people who use unregulated drugs.^22^ Conversely, changes in break and enters indicate an increase in spontaneous crimes and may be a consequence of coupling desire with the opportunity to execute said crimes.

Our results echo others’ observations that, *overall*, OPS/SCS did not contribute to increases in crimes.^8,11–13,23^ To contextualize our results, we consider the plausibility of three alternate explanations for our findings: the ‘honey-pot’ effect, increased policing, and changes in data collection.

The ‘honey-pot’ effect describes the phenomenon where an intervention attracts people and/or behaviours, and may explain why break and enters increased near OPS/SCS immediately following implementation.^5^ However, this phenomenon cannot explain why the initial increases in incidence did not persist. One reason may be that between March 2020 and August 2021 the city engaged in a massive displacement of people experiencing homelessness as part of its COVID-19 public health response. During this time, the city rented three large hotels (i.e., Bond, Edward Village, and Novotel) to provide temporary emergency shelter, effectively moving approximately 1000 people to other neighbourhoods. Meanwhile, the enforcement of stringent social distancing measures scaled down OPS/SCS services.^24^ The dispersing of clients to other neighbourhoods might explain the decline in spontaneous crimes. However, when measures were lifted, crime near OPS/SCS did not rebound despite a return to pre-pandemic rates of service use.^25^

Similarly, while increased police presence near OPS/SCS could explain some of the decline in spontaneous crimes, this presents a paradox. Previous studies noted police presence undermines the social acceptability of harm reduction interventions for people who use unregulated drugs.^26^ However, outside COVID-19 restrictions, there was no notable decline in client visits. Further, as far back as 2018, police budgets grew slower and the workforce *decreased*.^27,28^ The Toronto Police Service’s Mental Health and Addictions strategy launched in 2019 may help reconcile the shortages in the police workforce with decreases in crime reports.^29^ It is possible that by improving policing *quality*, crimes against persons, petty theft, and break and enters decreased with minimal effect on OPS/SCS patronage, and warrants investigation.

Finally, changes in the reporting or collecting of data may explain the decline in outcomes over time. However, in 2017, police services and Statistics Canada worked together to amend the definition of ‘founded’ criminal incidents to include events for which there was no credible evidence the incident *did not* take place.^30^ This victim-centred revision first came into effect 1 January 2018; and studies have associated it with 4 – 12% *increases* in counts of assault, petty theft, and fraud in subsequent years.^31^ While it is possible the public reported fewer crimes against persons near OPS/SCS – this seems unlikely when looking at city-wide reporting patterns.^32^

Nevertheless, like Hall et al., 2024, our analysis also found crime did increase near some OPS/SCS and neighbourhoods with multiple OPS/SCS (e.g., Moss Park and Church-Yonge) were most negatively impacted.^13^ The agglomeration of services, combined with unique geographic features may explain some of the differences observed. For example, the Moss Park neighbourhood boundaries include or are within 400m of Moss Park OPS, Fred Victor Centre, and Regent Park community health centre. Moss Park also includes housing and emergency shelter options for people experiencing homelessness. An influx of new people using OPS/SCS may have constrained existing resources and created tensions with incumbents resulting in an increase in assault reports. Meanwhile, St. Stephen’s is situated in Kensington Market, a neighbourhood full of small shops where opportunities to break and enter are higher than elsewhere in the city. Efforts to minimize break and enters (e.g., modernizing doorways and security) as well as declines in event reportings to prevent increases to insurance payments may *partially* explain the downward trend.

### Limitations

We did not include all OPS/SCS operating during the observation period. We excluded Casey House and shelters that provided services to residents only. Casey House became the first hospital to offer supervised consumption services to inpatients in August 2021, and expanded services to outpatients in April 2022. Because of differences in clientele, scope, duration of operation, and proximity to other sites – we excluded Casey House to minimize potentially biasing our analyses. Similarly, we did not include shelters which provided overdose prevention services to their residents, including the three temporary housing units implemented during COVID-19 social distancing efforts, as these are qualitatively different from OPS/SCS open to the public.

Further, we were not able to account for time-varying differential changes. For example, the observation period overlapped with significant upheaval in the housing market which may have contributed to differences in the degree of transient people experiencing homelessness in treated versus donor pool neighbourhoods. Owing to data availability, this remains a potential source of bias in our results.

Finally, we did not investigate the effects of OPS/SCS on public drug use, needle and syringe debris, graffiti or public defecation – concerns repeatedly mentioned by opponents of OPS/SCS.^33^ While we explored the possibility of including 311 calls in our study, we determined the quality of data insufficient and prone to reporting bias.

### Strengths

To our knowledge, this was the first study to evaluate the effects of multiple OPS/SCS implemented across a large metropolitan area. Owing to the number of sites and time elapsed since their implementation, we were sufficiently powered to examine the effects of OPS/SCS on a variety of measures separately, over time, and within proximity to sites. By pooling effect estimates by site and neighbourhood, we reduced the potential for unmeasured contemporaneous interventions biasing our results. Our synthetic controls were well matched to treated neighbourhoods (see reported root mean square prediction errors values in Supplement). We limited the risk of overfitting our models and achieving spurious pre-treatment fit by restricting analysis to three years.^34^ While imperfect matching between treated and synthetic controls may bias results, we combined traditional synthetic control methods with ITS to compensate for small differences in trends between treatment arms. By reporting the pooled and neighbourhood-specific synthetic control outputs, the effects of imperfect matching were made transparent, demonstrated to be minimal, and adequately managed using controlled interrupted time series methods.^35^ Further, by using exclusively open access data, and providing our code, we minimized the possibility for data preparation or analytic errors and increased the transparency of our work. Lastly, we considered other explanations for our observed effects and found the available evidence did not support these alternate narratives.

## PUBLIC HEALTH IMPACTS

Our analysis revealed the implementation of Toronto’s OPS/SCS had nuanced site- and neighbourhood-specific effects. Overall, the interventions’ impacts were neutral to positive over time. This does not negate the increases in spontaneous crimes in some communities, but rather suggests efforts to improve crime while encouraging use of OPS/SCS are possible. Still, local communities may perceive OPS/SCS as harmful, associating them with disorder and increased drug use in public spaces.^36^ This tension needs to be addressed to ensure the long term acceptability and utility of OPS/SCS. Efforts to establish relationships with local community stakeholders and to work collectively through challenges can build goodwill and trust between OPS/SCS operators, clients and the public. Meanwhile, efforts to dismiss or negate apprehensions can undermine the public health intervention.

## Data Availability

All data produced are available online at

https://data.torontopolice.on.ca/search?layout=grid

https://www150.statcan.gc.ca/n1/en/catalogue/92-168-X

