## Supplement File for "Evaluating the effects of Toronto’s supervised consumption sites on local crime"

**Title**

Evaluating the effects of Toronto’s supervised consumption sites on crime: multiple baseline interrupted time series analyses with and without synthetic controls

**Authors**

Dimitra Panagiotoglou^1^, PhD; Jihoon Lim^1^, PhD; Geoff Ingram^1^, BA; Mariam El Sheikh^1^, MScPH; Imen Farhat^1^, MS; Xander Bjornsson^1^, BSc; Maximilian Schaefer^1^, MScPH

1 Department of Epidemiology, Biostatistics and Occupational Health, McGill University, Montreal, Québec, Canada

Supplemental Table 1. OPS/SCS location, neighbourhood and dates of implementation, pg. 2

Supplemental Table 2. Definitions of each outcome included, pg. 3

Supplemental Table 3. Synthetic control predictor weights, pg. 4

Supplemental Figure 1. Church-Yonge and synthetic control interrupted time series results, assault, robbery, and break and enters, pg. 6

Supplemental Figure 2. Kensington-Chinatown and synthetic control interrupted time series results, assault, break and enters, and robberies, pg. 7

Supplemental Figure 3. Moss Park and synthetic control interrupted time series results, assault, break and enters, and robberies, pg. 8

Supplemental Figure 4. Niagara and synthetic control interrupted time series results, assault, break and enters, and robberies, pg. 9

Supplemental Figure 5. North St. James and synthetic control interrupted time series results, assault, break and enters, and robberies, pg. 10

Supplemental Figure 6. Regent Park and synthetic control interrupted time series results, assault, break and enters, and robberies, pg. 11

Supplemental Figure 7. Roncesvalles and synthetic control interrupted time series results, assault, break and enters, and robberies, pg. 12

Supplemental Figure 8. South Parkdale and synthetic control interrupted time series results, assault, break and enters, and robberies, pg. 13

Supplemental Figure 9. South Riverdale and synthetic control interrupted time series results, assault, break and enters, and robberies, pg. 14

Supplemental Figure 10. Trinity-Bellwoods and synthetic control interrupted time series results, assault, break and enters, and robberies, pg. 15

Supplemental Figure 11. Waterfront communities and synthetic control interrupted time series results, assault, break and enters, and robberies, pg. 16

For neighbourhood Donor Weights and Characteristics, please see attached file
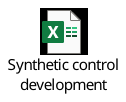

For additional distance figures (100m, 200m) please see attached file
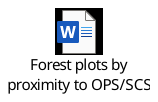

Supplemental Table 1. OPS/SCS location, neighbourhood and dates of implementation

| **Site** | **Neighborhood** | **Geo-Coordinates**  **(latitude, longitude)** | **Opened** | **Closed** |
| --- | --- | --- | --- | --- |
| Moss Park (OPS) | 73 – Moss Park | 43.65441862015198,  -79.37064612294073 | Aug 2017 | Jan 2018 |
| Moss Park (OPS) | 73 – Moss Park | 43.65420291365607,  -79.37196288883737 | Jan 2018 | May 2018 |
| Moss Park (OPS) | 73 – Moss Park | 43.654259892542775,  -79.36950796249596 | Jun 2018 |  |
| The Works (SCS) | 75 – Church-Yonge | 43.65666687196685,  -79.37952470339941 | Aug 2017 |  |
| South Riverdale Community Health Centre (SCS) | 70 – South Riverdale | 43.66121691864796,  -79.33924024717157 | Nov 2017 |  |
| Fred Victor Centre (SCS) | 73 – Moss Park  75 – Church-Yonge | 43.65354416730463,  -79.37289244572942 | Feb 2018 |  |
| Parkdale Queen West Community Health Centre (SCS) | 85 – South Parkdale  86 – Roncesvalles | 43.6419679071145,  -79.42942348805988 | Mar 2018 |  |
| Parkdale Supervised Consumption Service (OPS) | 77 – Waterfront-Communities  78 – Kensington  81 – Trinity-Bellwoods  82 – Niagara | 43.64661156407849,  -79.40405931100734 | Mar 2018 |  |
| Regent Park Community Health Centre (SCS) | 72 – Regent Park  73 – Moss Park | 43.65952867958244,  -79.36555506291515 | Apr 2018 |  |
| St. Stephens / KMOPS (OPS) | 78 – Kensington | 43.656560326757486,  -79.4028629707574 | Apr 2018 |  |
| Street Health (OPS) | 74 – North St. James Town  75 – Church-Yonge | 43.659455865049466,  -79.37024379204236 | Jun 2018 |  |

Supplemental Table 2. Definitions of each outcome included

We restricted analyses to founded crimes that occurred between 1 January 2014 and 30 June 2024; and fall under two types of offences within Canada’s Criminal Code: offences against rights of property (e.g., theft) and crimes against persons. Unlike other criminal systems, there is no formal distinction between ‘petty’ and ‘grand’ theft, except by way of penalty, using $5000 as the threshold. We considered bicycle theft and theft from vehicle as petty theft, motor vehicle theft and theft over $5000 as grand theft, and assault and robbery as crimes against persons given the presence of a victim.^1^ Break and enters are also offences against property but were not categorized as grand or petty theft.

| Outcome | Definition |
| --- | --- |
| Assault | All intentional direct or indirect application and attempt of force to another person. Assault also includes attempts or threats, by act or gesture, to apply force to another person; or causes other person to believe on reasonable grounds that they have, present ability to achieve their purpose. |
| Auto thefts | Limited to acts of taking another person’s vehicle. |
| Break and enters | Acts of entering places with the *intent* of committing indictable offenses (e.g., involving a dwelling house with no lawful excuse). |
| Robberies | Acts of taking property from a person or business using force or intimidation in the presence of a victim. |
| Thefts over $5000 | Stealing property in excess of $5000 (excluding auto theft). |
| Bicycle thefts | Any occurrence where theft of bicycle occurred. |
| Thefts from motor vehicles | Acts of stealing property from a motor vehicle. |
| Mental Health Act apprehensions | Include all police apprehensions of persons identified as a risk to themselves or others for the provision of medical care and up to 72 hours of observation. |

Supplemental Table 3. Synthetic control predictor weights

|  | **Church-Yonge** | | | | | | | | **Kensington-Chinatown** | | | | | | | | **Moss Park** | | | | | | | | **Niagara** | | | | |
| --- | --- | --- | --- | --- | --- | --- | --- | --- | --- | --- | --- | --- | --- | --- | --- | --- | --- | --- | --- | --- | --- | --- | --- | --- | --- | --- | --- | --- | --- |
| **Predictor** | **Assault** | | **B&E** | | | | **Robbery** | | **Assault** | | **B&E** | | | | **Robbery** | | **Assault** | | **B&E** | | | **Robbery** | | | **Assault** | | **B&E** | | **Robbery** |
| Total population | 0.01 | | 0.01 | | | | 0.08 | | 0 | | 0.16 | | | | 0.17 | | 0.01 | | 0.04 | | | 0.02 | | | 0.03 | | 0.15 | | 0.02 |
| Population density | 0.02 | | 0.08 | | | | 0.06 | | 0 | | 0.21 | | | | 0.17 | | 0.16 | | 0.01 | | | 0 | | | 0.09 | | 0.09 | | 0.28 |
| Female (%) | 0.01 | | 0.01 | | | | 0.01 | | 0.13 | | 0.21 | | | | 0.12 | | 0 | | 0.02 | | | 0 | | | 0.04 | | 0.13 | | 0 |
| Ages 0-14 (%) | 0.22 | | 0.02 | | | | 0.24 | | 0 | | 0.05 | | | | 0.11 | | 0.02 | | 0.08 | | | 0.04 | | | 0.03 | | 0.04 | | 0.02 |
| Ages 15-64 (%) | 0.37 | | 0.38 | | | | 0.43 | | 0.01 | | 0.1 | | | | 0.11 | | 0.12 | | 0.03 | | | 0.11 | | | 0.03 | | 0 | | 0.07 |
| Ages 65+ | 0.31 | | 0.21 | | | | 0.05 | | 0.13 | | 0.01 | | | | 0 | | 0.25 | | 0.06 | | | 0.21 | | | 0.19 | | 0.02 | | 0 |
| Average age | 0 | | 0.05 | | | | 0.01 | | 0.09 | | 0 | | | | 0 | | 0.11 | | 0.13 | | | 0 | | | 0.01 | | 0.08 | | 0 |
| Household size | 0.01 | | 0.01 | | | | 0.09 | | 0 | | 0.19 | | | | 0.04 | | 0.07 | | 0.42 | | | 0.26 | | | 0 | | 0.1 | | 0.02 |
| Median income | 0.01 | | 0.01 | | | | 0 | | 0.01 | | 0 | | | | 0.14 | | 0.03 | | 0.06 | | | 0.16 | | | 0.05 | | 0 | | 0.01 |
| Low income measure - After tax | 0.01 | | 0.11 | | | | 0 | | 0.02 | | 0 | | | | 0.02 | | 0.07 | | 0.1 | | | 0 | | | 0.36 | | 0.18 | | 0.38 |
| One parent household (%) | 0 | | 0.02 | | | | 0 | | 0.1 | | 0.01 | | | | 0.03 | | 0.02 | | 0.03 | | | 0 | | | 0 | | 0.01 | | 0 |
| Immigrant (%) | 0.01 | | 0.01 | | | | 0.01 | | 0 | | 0.01 | | | | 0 | | 0 | | 0.01 | | | 0.05 | | | 0.03 | | 0.01 | | 0.01 |
| Visible minority (%) | 0.01 | | 0.08 | | | | 0 | | 0.28 | | 0 | | | | 0.09 | | 0.03 | | 0.02 | | | 0 | | | 0.07 | | 0.05 | | 0.05 |
| No education (%) | 0 | | 0 | | | | 0 | | 0.17 | | 0.01 | | | | 0 | | 0.07 | | 0 | | | 0 | | | 0.02 | | 0.02 | | 0.03 |
| Unemployment rate (%) | 0.01 | | 0 | | | | 0 | | 0.05 | | 0.04 | | | | 0 | | 0.05 | | 0.02 | | | 0.14 | | | 0.05 | | 0.12 | | 0.11 |
|  | | **Roncesvalles** | | | | | | | **South Parkdale** | | | | | | | | **South Riverdale** | | | | | | | | **Trinity-Bellwoods** | | | | |
| **Predictor** | | **Assault** | | **B&E** | | **Robbery** | | | **Assault** | | | **B&E** | | **Robbery** | | | **Assault** | | | **B&E** | | | **Robbery** | | **Assault** | **B&E** | | **Robbery** | |
| Total population | | 0.16 | | 0.15 | | 0.07 | | | 0.06 | | | 0.01 | | 0 | | | 0.02 | | | 0.15 | | | 0.05 | | 0.17 | 0.1 | | 0.02 | |
| Population density | | 0.08 | | 0.02 | | 0.06 | | | 0.1 | | | 0 | | 0 | | | 0.01 | | | 0.12 | | | 0 | | 0.08 | 0.07 | | 0 | |
| Female (%) | | 0.14 | | 0.15 | | 0.25 | | | 0.1 | | | 0.03 | | 0.19 | | | 0.03 | | | 0.09 | | | 0.16 | | 0.1 | 0.01 | | 0.11 | |
| Ages 0-14 (%) | | 0.01 | | 0.07 | | 0 | | | 0.05 | | | 0.09 | | 0.08 | | | 0.19 | | | 0 | | | 0.07 | | 0.15 | 0.07 | | 0.09 | |
| Ages 15-64 (%) | | 0.11 | | 0.06 | | 0 | | | 0 | | | 0.03 | | 0.07 | | | 0.33 | | | 0.02 | | | 0.14 | | 0.01 | 0.04 | | 0 | |
| Ages 65+ | | 0.01 | | 0.07 | | 0.05 | | | 0.05 | | | 0 | | 0.03 | | | 0.31 | | | 0 | | | 0.02 | | 0.02 | 0.09 | | 0.05 | |
| Average age | | 0.12 | | 0.2 | | 0.1 | | | 0.03 | | | 0.11 | | 0.09 | | | 0.01 | | | 0.08 | | | 0.13 | | 0.05 | 0.07 | | 0 | |
| Household size | | 0 | | 0 | | 0.14 | | | 0.01 | | | 0.03 | | 0 | | | 0 | | | 0.01 | | | 0.09 | | 0.04 | 0.01 | | 0.18 | |
| Median income | | 0.02 | | 0 | | 0.02 | | | 0.06 | | | 0.02 | | 0 | | | 0.02 | | | 0.01 | | | 0.04 | | 0 | 0.1 | | 0.21 | |
| Low income measure - After tax | | 0.05 | | 0 | | 0.04 | | | 0.2 | | | 0.02 | | 0 | | | 0.04 | | | 0.16 | | | 0.01 | | 0 | 0.01 | | 0.07 | |
| One parent household (%) | | 0.05 | | 0.2 | | 0.09 | | | 0.1 | | | 0.31 | | 0.11 | | | 0.01 | | | 0.19 | | | 0 | | 0.01 | 0.18 | | 0.08 | |
| Immigrant (%) | | 0.04 | | 0 | | 0 | | | 0.02 | | | 0.02 | | 0.02 | | | 0 | | | 0.04 | | | 0.03 | | 0.04 | 0.06 | | 0 | |
| Visible minority (%) | | 0.04 | | 0 | | 0.12 | | | 0.02 | | | 0.01 | | 0.03 | | | 0.04 | | | 0.13 | | | 0.26 | | 0.02 | 0.09 | | 0 | |
| No education (%) | | 0.02 | | 0 | | 0.06 | | | 0.07 | | | 0.14 | | 0.17 | | | 0 | | | 0 | | | 0 | | 0.18 | 0 | | 0.16 | |
| Unemployment rate (%) | | 0.14 | | 0.07 | | 0 | | | 0.14 | | | 0.17 | | 0.22 | | | 0 | | | 0.01 | | | 0 | | 0.12 | 0.1 | | 0.04 | |
|  | | **North St. James** | | | | | | **Regent Park** | | | | | | | | **Waterfront Communities** | | | | | | | |  |  |  |  |  |  |
| **Predictor** | | **Assault** | | **B&E** | **Robbery** | | | **Assault** | | **B&E** | | | **Robbery** | | | **Assault** | | **B&E** | | | **Robbery** | | |  |  |  |  |  |  |
| Total population | | 0.05 | | 0.01 | 0.09 | | | 0.1 | | 0.06 | | | 0.11 | | | 0.03 | | 0.01 | | | 0.09 | | |  |  |  |  |  |  |
| Population density | | 0.07 | | 0 | 0 | | | 0 | | 0 | | | 0 | | | 0.01 | | 0 | | | 0.06 | | |  |  |  |  |  |  |
| Female (%) | | 0.12 | | 0.23 | 0.03 | | | 0.15 | | 0.09 | | | 0.09 | | | 0.01 | | 0.03 | | | 0.01 | | |  |  |  |  |  |  |
| Ages 0-14 (%) | | 0.14 | | 0.15 | 0 | | | 0.03 | | 0.22 | | | 0.06 | | | 0.22 | | 0.24 | | | 0.11 | | |  |  |  |  |  |  |
| Ages 15-64 (%) | | 0.03 | | 0.19 | 0 | | | 0.08 | | 0.11 | | | 0.13 | | | 0.35 | | 0.39 | | | 0.05 | | |  |  |  |  |  |  |
| Ages 65+ | | 0.03 | | 0.11 | 0.05 | | | 0.05 | | 0.05 | | | 0.14 | | | 0.29 | | 0.31 | | | 0.04 | | |  |  |  |  |  |  |
| Average age | | 0.01 | | 0.01 | 0.13 | | | 0.07 | | 0.08 | | | 0.05 | | | 0.02 | | 0 | | | 0.1 | | |  |  |  |  |  |  |
| Household size | | 0 | | 0 | 0 | | | 0.01 | | 0.02 | | | 0.06 | | | 0.01 | | 0 | | | 0.13 | | |  |  |  |  |  |  |
| Median income | | 0.07 | | 0.01 | 0.05 | | | 0.01 | | 0.02 | | | 0.11 | | | 0.01 | | 0 | | | 0.01 | | |  |  |  |  |  |  |
| Low income measure - After tax | | 0.03 | | 0 | 0.26 | | | 0 | | 0 | | | 0 | | | 0.01 | | 0 | | | 0.1 | | |  |  |  |  |  |  |
| One parent household (%) | | 0.17 | | 0.08 | 0.12 | | | 0.21 | | 0.26 | | | 0.11 | | | 0.01 | | 0 | | | 0.07 | | |  |  |  |  |  |  |
| Immigrant (%) | | 0.01 | | 0.1 | 0.09 | | | 0.03 | | 0 | | | 0.1 | | | 0.01 | | 0 | | | 0.04 | | |  |  |  |  |  |  |
| Visible minority (%) | | 0.11 | | 0 | 0.06 | | | 0.04 | | 0.05 | | | 0 | | | 0.01 | | 0 | | | 0 | | |  |  |  |  |  |  |
| No education (%) | | 0.1 | | 0.01 | 0.04 | | | 0.17 | | 0.03 | | | 0.01 | | | 0 | | 0 | | | 0.17 | | |  |  |  |  |  |  |
| Unemployment rate (%) | | 0.05 | | 0.09 | 0.07 | | | 0.03 | | 0.01 | | | 0.01 | | | 0.01 | | 0 | | | 0.02 | | |  |  |  |  |  |  |

Supplemental Figure 1. Church-Yonge Street and Synthetic Control interrupted time series results, assault, break and enters, and robberies

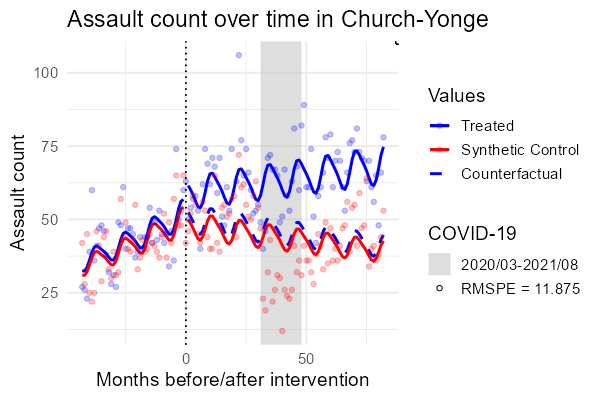

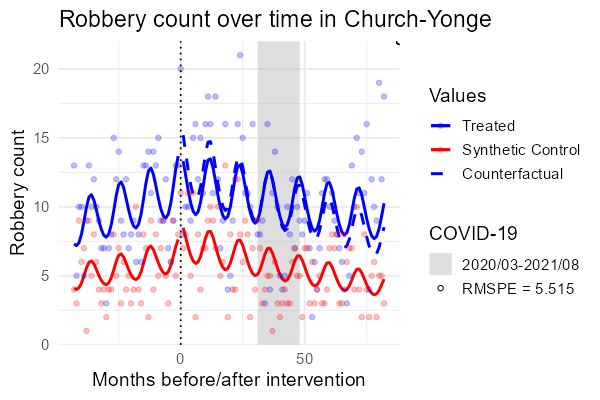

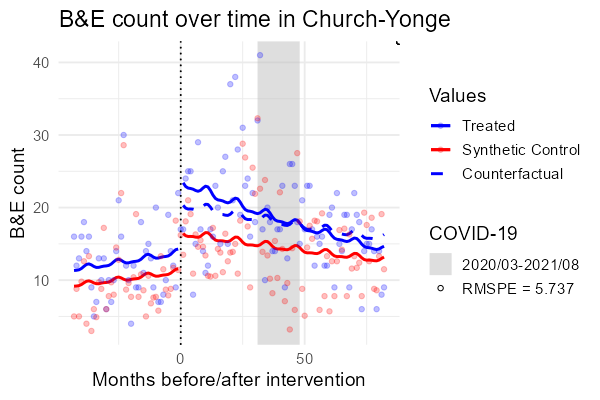

Supplemental Figure 2. Kensington-Chinatown and Synthetic Control interrupted time series results, assault, break and enters, and robberies

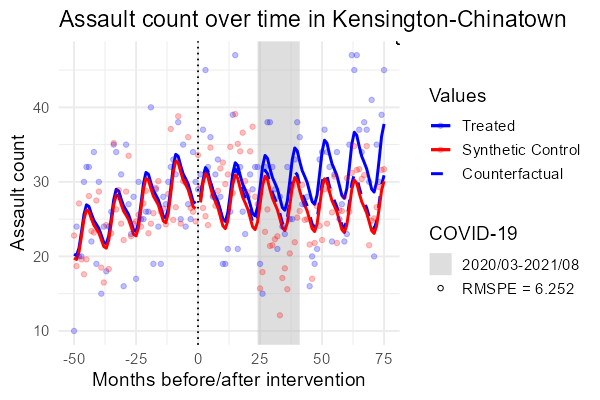

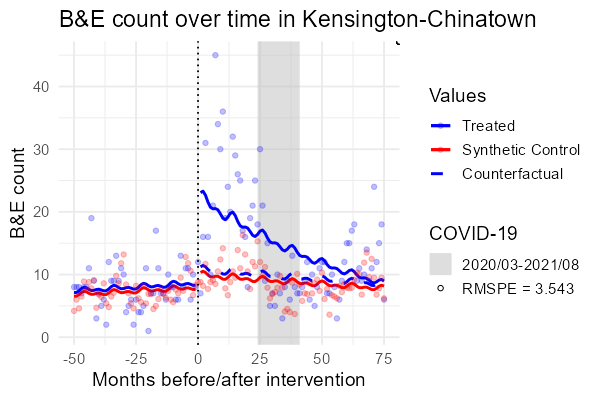

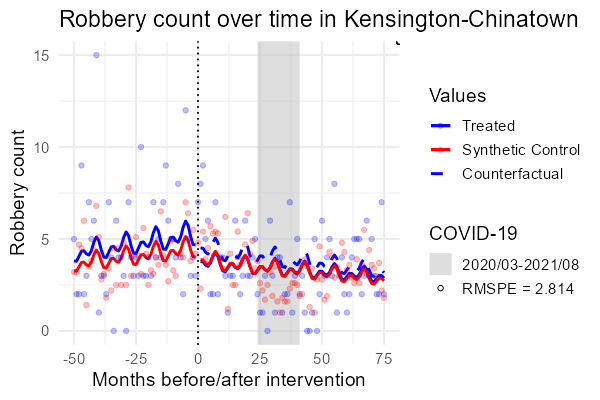

Supplemental Figure 3. Moss Park and Synthetic Control interrupted time series results, assault, break and enters, and robberies

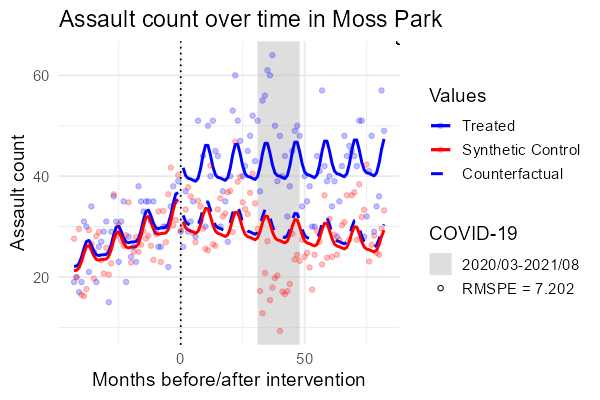

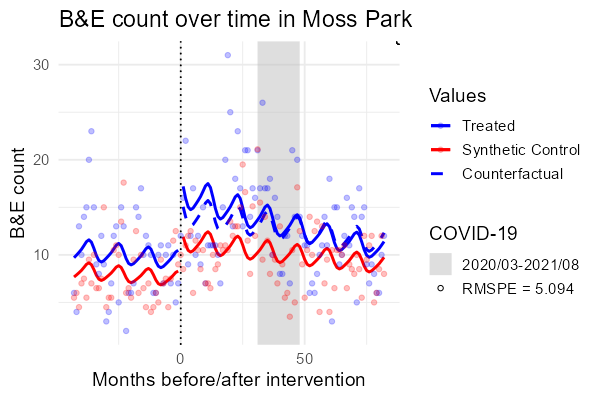

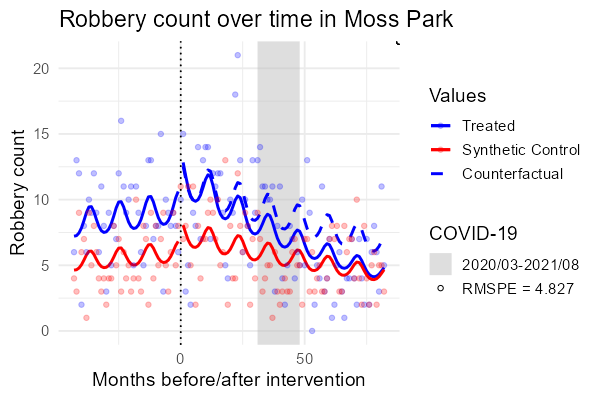

Supplemental Figure 4. Niagara and synthetic control interrupted time series results, assault, break and enters, and robberies

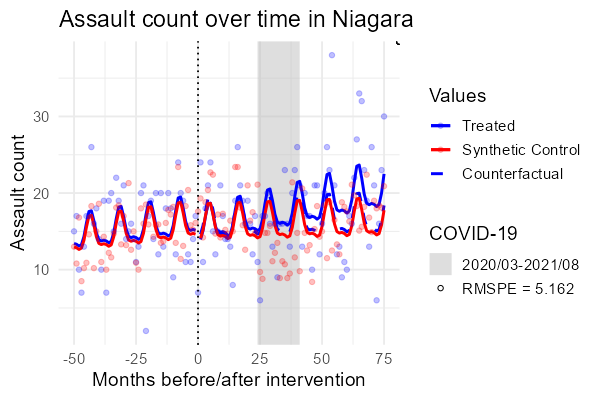

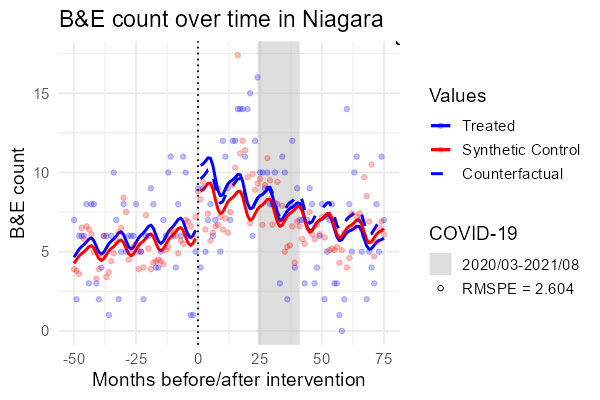

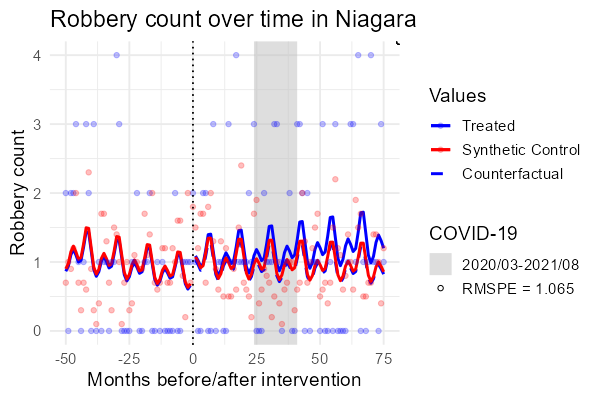

Supplemental Figure 5. North St. James and synthetic control interrupted time series results, assault, break and enters, and robberies

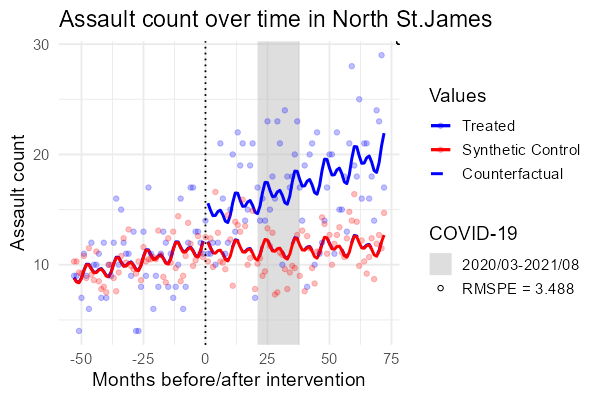

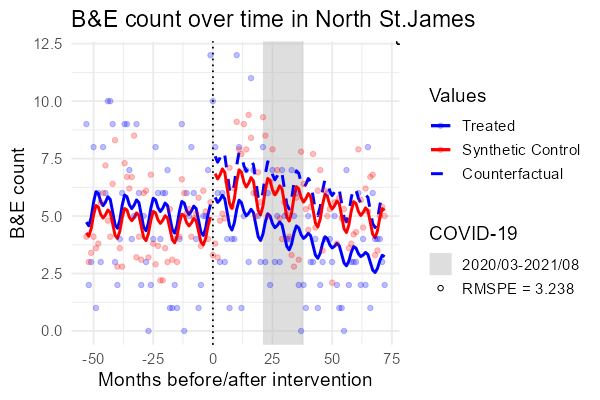

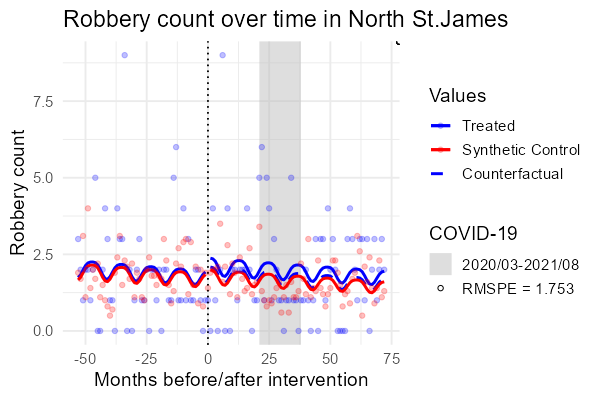

Supplemental Figure 6. Regent Park and synthetic control interrupted time series results, assault, break and enters, and robberies

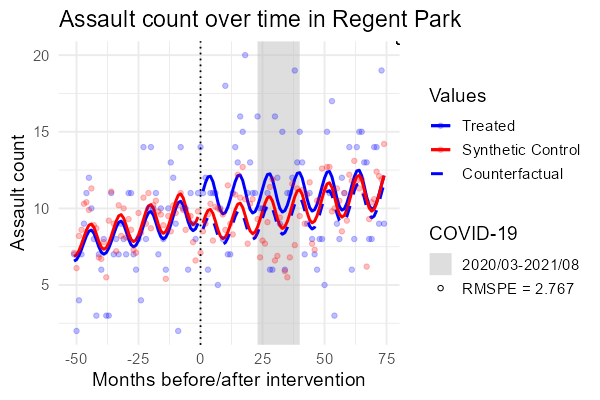

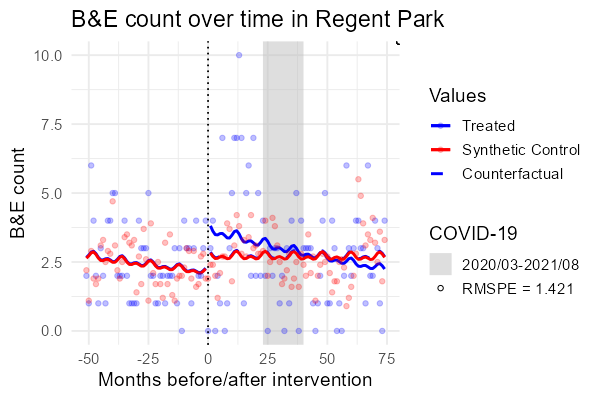

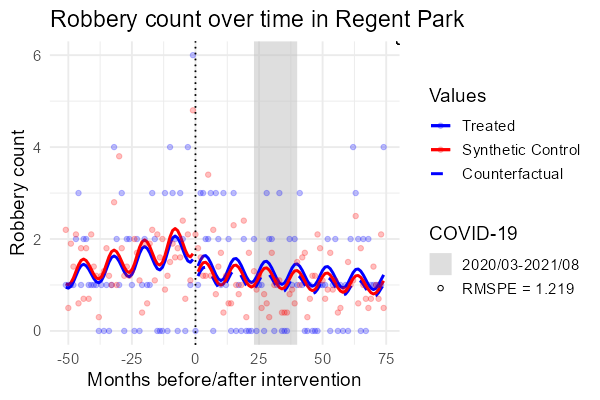

Supplemental Figure 7. Roncesvalles and synthetic control interrupted time series results, assault, break and enters, and robberies

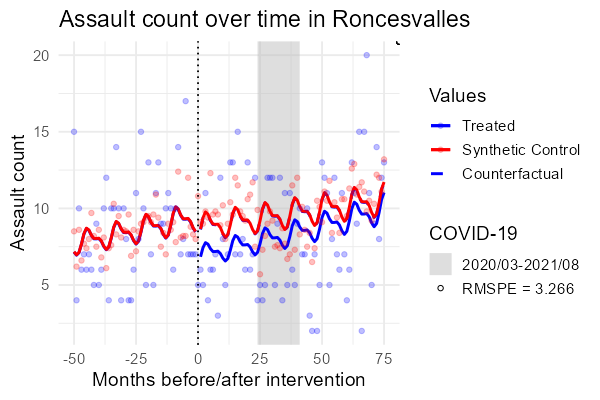

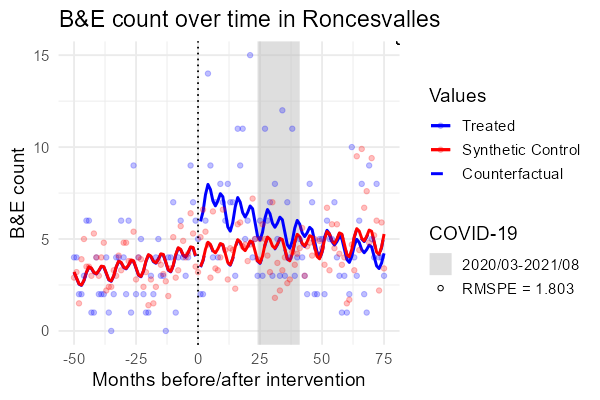

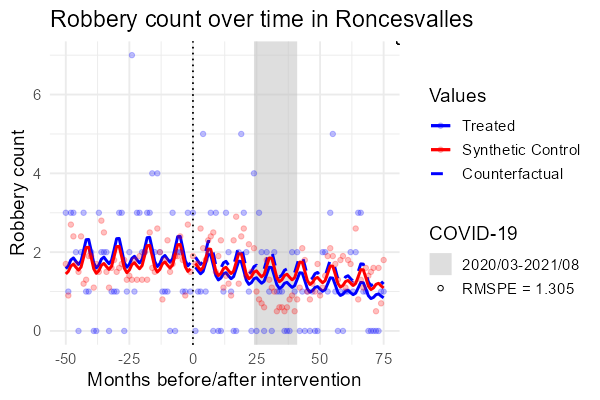

Supplemental Figure 8. South Parkdale and synthetic control interrupted time series results, assault, break and enters, and robberies
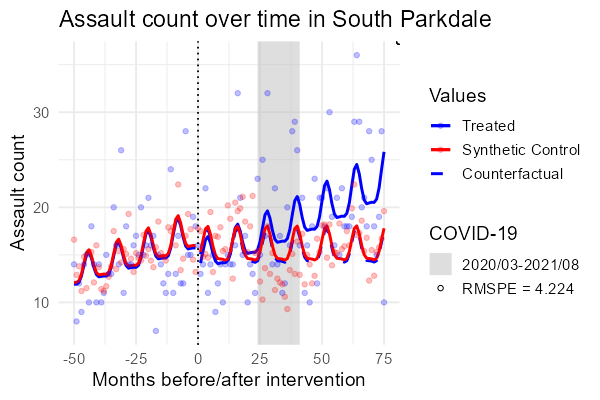

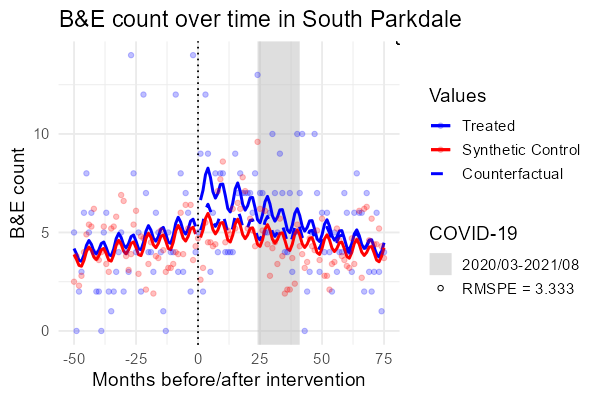

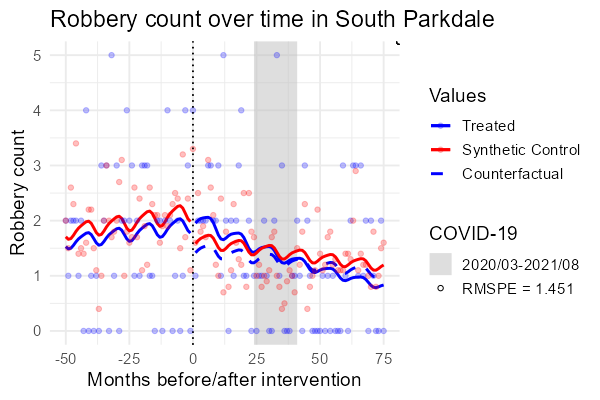

Supplemental Figure 9. South Riverdale and synthetic control interrupted time series results, assault, break and enters, and robberies

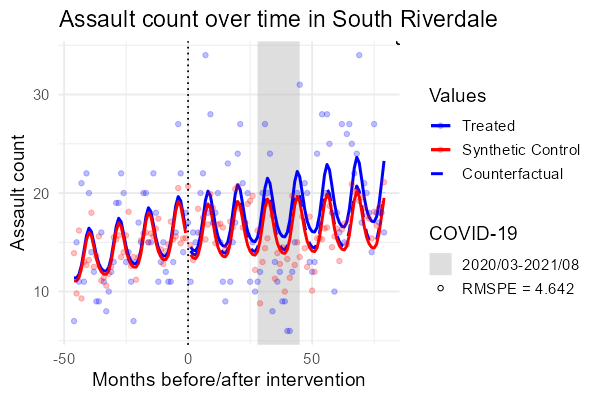

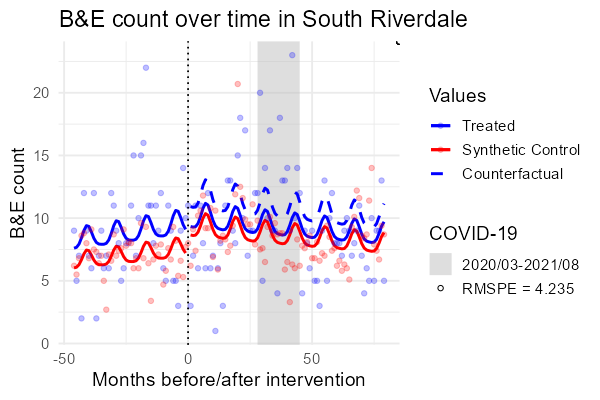

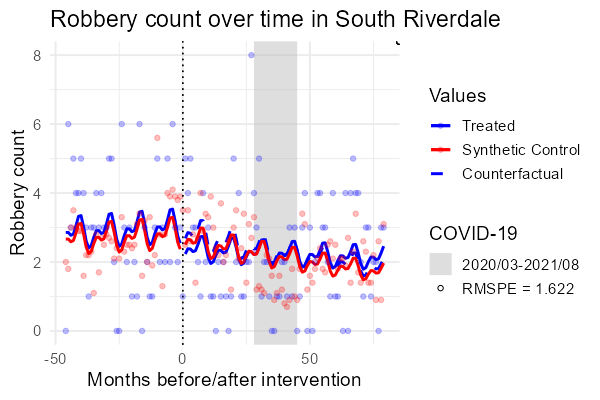

Supplemental Figure 10. Trinity-Bellwoods and synthetic control interrupted time series results, assault, break and enters, and robberies

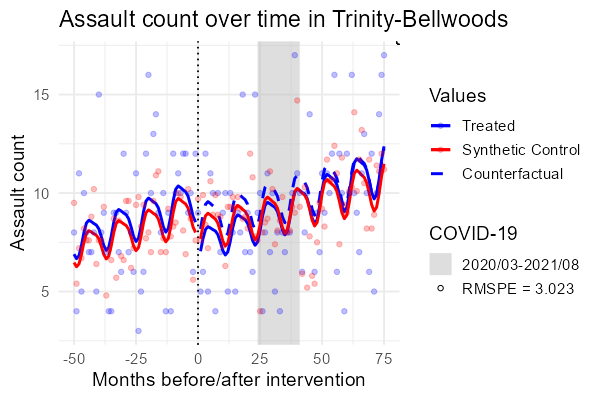

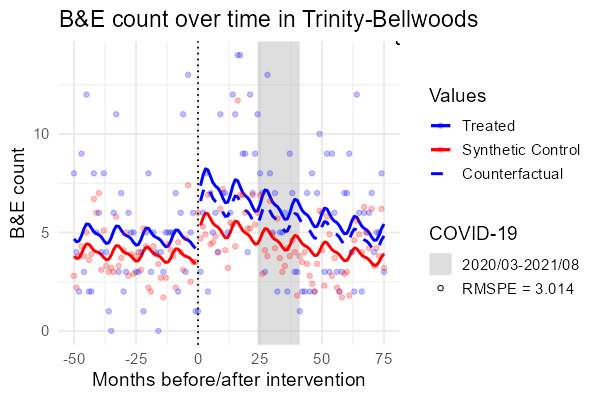

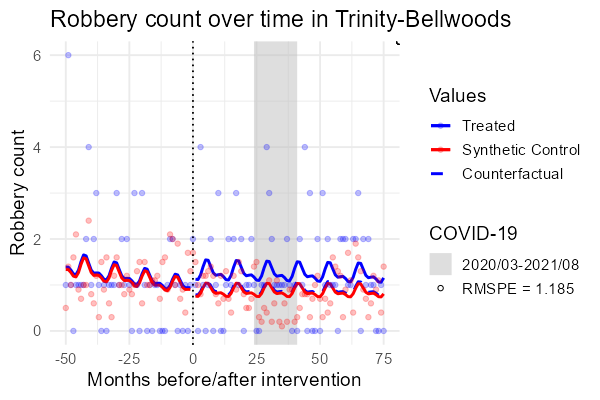

Supplemental Figure 11. Waterfront communities and synthetic control interrupted time series results, assault, break and enters, and robberies

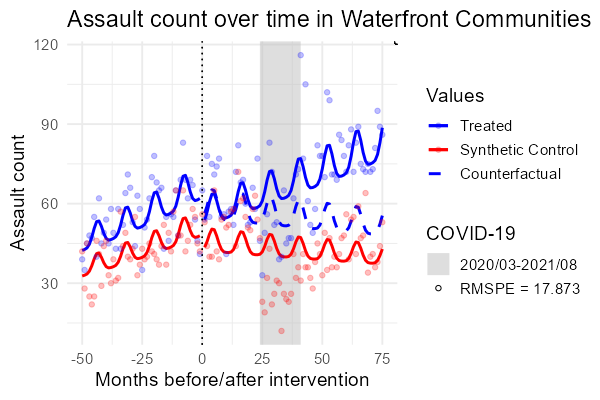

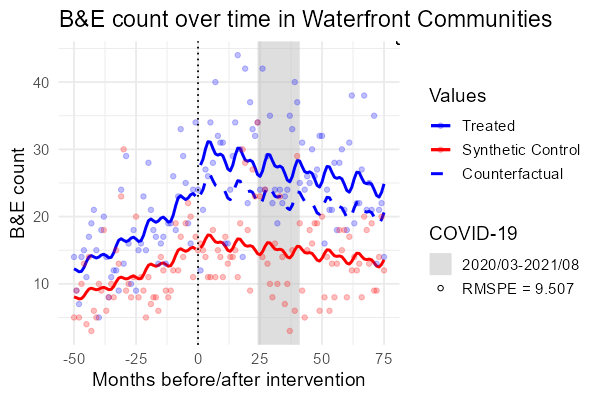

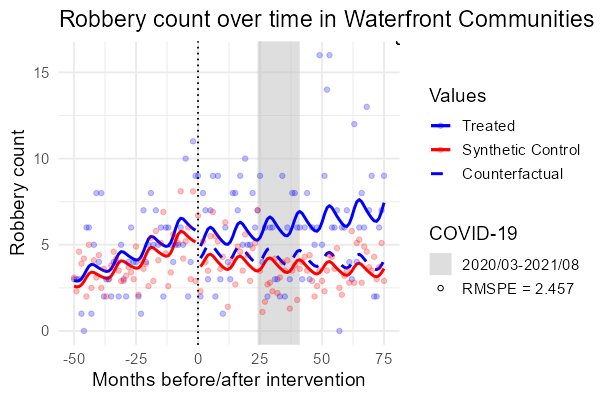

1.	Statistics Canada. Classification of common offence. Accessed 26 September 2024, <https://www23.statcan.gc.ca/imdb/p3VD.pl?Function=getVD&TVD=257740&CVD=257742&CPV=1.1&CST=01012015&CLV=1&MLV=3&D=1>
